# Prolonged viral shedding as a marker of severity in respiratory syncytial virus bronchiolitis

**DOI:** 10.1101/2024.08.29.24312713

**Authors:** Raí André Silva Watanabe, Filipe Nishiyama, Larissa Lyra, Bruno Sanchez Camargo, Luciano Kleber de Souza Luna, Danielle Dias Conte, Ana Helena Perosa, Nancy Cristina Junqueira Bellei

## Abstract

**Background and Objectives:** Respiratory syncytial virus (RSV) is responsible for most cases of acute viral bronchiolitis (AVB) in childhood. The association of factors such as RSV subtype, viral load, and viral coinfection with severe disease is controversial. The objective is to describe the viral load dynamics of RSV in children under 2 years with AVB, including the first viral load, peak viral load, viral decay, and any possible association with severe disease.

**Methods:** 73 inpatients with AVB and confirmed RSV infection were included. Viral load was obtained through nasal swab samples daily during hospitalization and weekly after discharge until absence of detection.

**Results:** 44 of the patients were male, the mean age was 5.76 months, and comorbidities were found in 15 (20.5%) of the patients. 54 (74%) of the patients had the peak viral load between the 4th and the 7th day of symptom onset. The mean duration of viral detection was 12.5 days. There was no association between the first and peak viral load with markers of severe disease. However, association was found between viral persistence exceeding 10 days and the following factors: longer hospitalization period (p=0.009), length of ventilation support (p=0.044), length of invasive mechanical ventilation (p=0.035), prolonged requirement of nutritional support (p=0.038), and a longer course of antibiotic treatment (p=0.024). Age was inversely correlated with most of the severity outcomes.

**Conclusion:** Despite the lack of association between RSV viral load values and disease severity, prolonged viral shedding was associated with several adverse outcomes, which can contribute to a better understanding of RSV disease, particularly as several interventions are anticipated to become available in the coming years.

## Introduction

### Prolonged viral shedding as a marker of severity in respiratory syncytial virus bronchiolitis

Respiratory syncytial virus (RSV) is the major cause of lower respiratory tract infections and accounts for up to 80% of acute viral bronchiolitis (AVB) in childhood^1,2,3^. Nearly every child under two years of age is infected with the virus^1^, and 2-3% require hospitalization^3^. Recent data from the United States indicates that RSV is responsible for 9.3% of all hospitalizations among the pediatric population with an estimate burden of 1.2 billion dollars each year^4^. Two multi-site, international case-control studies about pneumonia etiology in children under 5 years of age were conducted in the last decade and RSV had the highest etiological fraction on the PERCH study and the second highest on the GABRIEL study^5,6^.

Up to 30% of AVB cases involve the detection of multiple viruses, and most cases occur within the first six months of life^3,7^. The risk factors for severe AVB includes lower age (specially below 3 months), prematurity, bronchopulmonary dysplasia, congenital heart disease, immunodeficiency, genetic disorders, neurological and neuromuscular disease, tobacco exposure during pregnancy, lack of breastfeeding and male sex ^3,7^.

Studies on RSV shedding, and clinical severity vary in methodology, age groups studied, diagnostic criteria, risk factors considered, and definitions of outcomes. Furthermore, there are variations in the timing of viral load measurements and the methods used for quantification^8–13^.

New interventions for managing RSV infections, such as vaccines and monoclonal antibodies, have recently been approved^14^. To assess their impact on the dynamics of RSV-associated AVB, it is essential to increase the understanding of RSV viral load.

The study’s objective was to characterize the dynamics of RSV viral load from the time of hospital admission to the point of viral shedding, encompassing both inpatients and outpatients. Additionally, the study intended to investigate potential associations between viral load, the RSV subgroup type, the detection of multiple viruses, and patient characteristics (such as age, gender, and comorbidities) and their impact on the severity of clinical outcomes.

## Materials and Methods

### Study Design

The study aimed to characterize the dynamics of RSV viral load, tracking it from hospital admission to the point of final viral shedding, in both inpatients and outpatients. In addition, the study sought to investigate potential associations between viral load, the RSV subgroup type, the presence of multiple viruses excluding severe acute respiratory syndrome coronavirus 2 (SARS-CoV-2), and patient characteristics (such as age, gender, and comorbidities) in relation to the severity of clinical outcomes.

The inclusion criteria for the study were as follows:

1. Hospitalized patients under 2 years of age diagnosed with acute viral bronchiolitis and not expected to be discharged within the next 24 hours.
2. Initial assessment confirming RSV infection through reverse transcription – quantitative Polymerase Chain Reaction (RT-qPCR).

The exclusion criteria were:

1. Loss of follow-up before a viral load sample with no RSV detection.
2. Patients with less than 3 collected samples.
3. Detection of SARS-CoV-2 through RT-qPCR for a child included in the pandemic period.

### Data collection

The study utilized a convenience sample of children admitted to the hospital’s pediatric service with a clinical diagnosis of AVB. These children underwent nasopharyngeal swabs to detect RSV through RT-qPCR. The diagnosis of AVB was made by the attending pediatric physician in accordance with the guidelines of the American Academy of Pediatrics^15^. Nasopharyngeal swab samples were obtained by the team coordinator, trained resident physicians, or the on-duty physiotherapist for laboratory confirmation of RSV infection. Samples were collected daily during the period of the infant’s hospitalization and weekly after hospital discharge, with the collection stopping when no further RSV was detected in two consecutive samples. The day of symptom onset was defined as the first respiratory or systemic symptom of RSV infection: fever, cough, runny nose, nasal obstruction, cyanosis, apnea, respiratory distress or wheezing.

### Laboratory Methods

Each collected swab was preserved in 2.0 mL of sterile lactated Ringer’s solution and immediately transported to the virology laboratory for RSV molecular screening on the same day or stored at -80°C until investigation. The RNA of swab samples was purified using the QIAamp Viral RNA Mini Kit (Qiagen, Hilden, Germany) according to the manufacturer’s instructions. Molecular detection of RSV was performed by a duplex quantitative RT-qPCR with TaqMan Fast Virus 1-Step Master Mix reagents (Applied Biosystems, Vilnius, Lithuania). The 20 µL of reaction volume contained 1 µM of each primer and 250 nM of the probe for RSV detection (labeled with FAM dye), aimed at the Matrix gene^16^, and 250 nM of each primer and probe (labeled with VIC dye) for human ribonuclease P gene detection as an internal control^17^. Thermocycling conditions were performed on a 7500 Real-Time PCR System (Applied Biosystems), as follow: 50°C for 2 min., 95°C for 20 sec., followed by 45 cycles of 95°C for 15 sec., and 55°C for 30 sec. (data collection). The cycle threshold of 35 was adopted as a cut-off for the human ribonuclease P and 38 for the RSV RT-qPCR.

### Viral load analysis

The viral load was measured from a standard curve generated as described elsewhere^18^, with an efficiency of 100.52%. The viral load was expressed in Log10 RNA copies/mL.

### RSV Subtype

The protocol of RSV subtyping was performed by Fiocruz laboratory with the sample of the highest viral load of each patient^19^.

### Influenza and Rhinovirus detection

An in-house real-time PCR was performed to detect Influenza A, B and rhinovirus with AgPath-ID™ One-Step RT-PCR Reagents (ThermoFischer) according to the manufacturer’s instructions. The primers and probes used for each virus detection have been published elsewhere^20,21^. A positive result was considered with a Ct < 40.

### Statistics

Sample size calculation was performed using G Power 3.1.9.7, based on an effect size of 0.4, power of 80%, alfa error probability of 5% using a linear regression model with 7 predictor variables. The number of patients to achieve statistical significance was 44. Data was stored in Microsoft excel 365 sheets. Statistical analysis was obtained using Jamovi (version 2.3.13) with alfa error probability of 5% and confidence interval of 95%. Graphs were built with Microsoft excel 365 and SPSS 26^th^ edition.

Continuous variables were described as mean with standard deviation and median with interquartile range, and categorical variables as frequency rates. The 10-day cutoff for viral persistence as a categorical variable was based on the median number obtained as viral persistence.

Results were considered statistically significant when *p* value was below 0,05. Multiple linear regression was used accordingly. R^2^ was used to report the adequation of the model to the data and for effect size. Two models were performed, according to the equation:

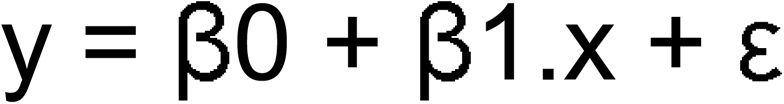

y = dependent variable

x = covariates

β0 = intercept

β1 = angular coefficient

ε = error

Model A: Dependent variables: (1) viral persistence; (2) peak viral load; (3) first viral load. Covariates: (1) age; (2) sex; (3) RSV subtype; (4) comorbidities; (5) rhinovirus.

Model B: Dependent variables (days of each outcome): (1) hospitalization; (2) fever; (3) use of oxygen; (4) ICU; (5) any ventilation support; (6) IMV; (7) additional nutrition support; (8) use of antibiotics. Covariates: (1) age; (2) peak viral load; (3) viral persistence over 10 days; (4) sex; (5) RSV subtype; (6) comorbidities; (7) rhinovirus.

## Results

Out of a total of 191 hospitalized patients under 2 years with AVB who were tested for RSV infection, 112 (63,9%) were positive for RSV and 73 patients were enrolled in the study. The main reasons of the exclusion for the 48 patients that had RSV detection were: (1) discharge under 24 hours; (2) hospital transfer before 3 samples collected; (3) refusal to participate. One patient had SARS-CoV-2 detection. In 2019, 40 patients were included, and in 2021, 33 patients were included. A total of 548 nasopharyngeal samples were collected from the enrolled patients, averaging 7.5 samples per patient. Out of these, 496 (90.5%) were obtained during the hospitalization period and were collected daily. Before discharge, 42 patients (57.5%) exhibited complete viral clearance.

### Characteristics of study population

The characteristics of the study population are outlined in Table 1. Comorbidities were observed in 9 patients with prematurity (12.3%), 3 patients with cardiac and neurological diseases (4.1% each), and one patient with one of the following: immunodeficiency, bronchopulmonary dysplasia, hematologic disease, renal disease, and genetic syndrome. Two of the patients had more than one comorbidity. None of the premature infants had a gestational age below 30 weeks (ranging from 30 to 36 weeks), and only one received palivizumab. The patient who received palivizumab had down syndrome, bronchopulmonary dysplasia, and cardiac disease (patient number 38 from figure 2).

Recurrent wheezing was not considered a comorbidity, and 8 patients in the 2019 cohort had experienced at least one previous wheezing episode. Among the 12 cases with rhinovirus co-infection, lower viral loads and shorter persistence of rhinovirus was observed compared to RSV values of the same patient. The Ct values for rhinovirus RNA detection ranged from 28 to 39.

### Viral Load Analysis

All patients had their first nasopharyngeal swab samples collected on the first or second day of hospitalization. Upon admission, the mean day of symptom onset was 5.11 ± 2.54, with a median of 5 (ranging from 4 to 6). Three caregivers reported symptoms lasting for more than 12 days at the time of admission.

The initial viral load in the first sample ranged from 4 to 8.01 log10 copies/ml, with a mean of 6.03 ± 1.10 and a median of 6.34 (ranging from 5.12 to 6.79). Peak viral load varied between 4.14 and 8.01 log10 copies/ml, with a mean of 6.3 ± 0.98 and a median of 6.49 (ranging from 5.67 to 6.92). In 40 patients (54.8%), the first viral load was identical to the peak viral load. The time to reach the peak viral load after the onset of symptoms is detailed in Table 2.

Individual viral load decay can be observed in Figures 1 to 4, while the distribution of viral loads for all patients based on the onset of symptoms is illustrated in Figure 5.

**Figure 1.**
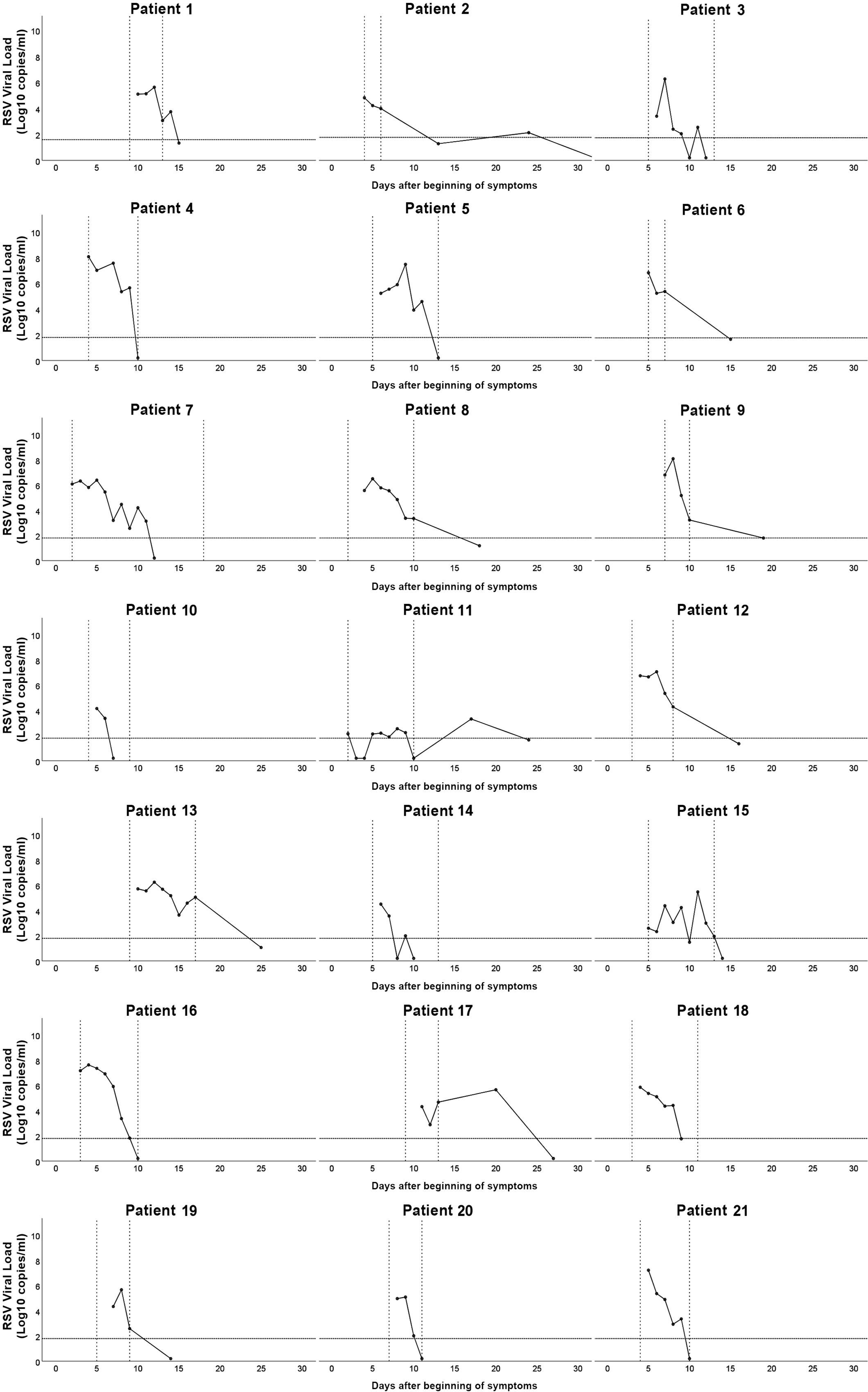
**RSV viral load decay, patients 1 to 21. Horizontal line corresponds to the cut of viral detection. Vertical lines indicate period of hospitalization.**

**Figure 2.**
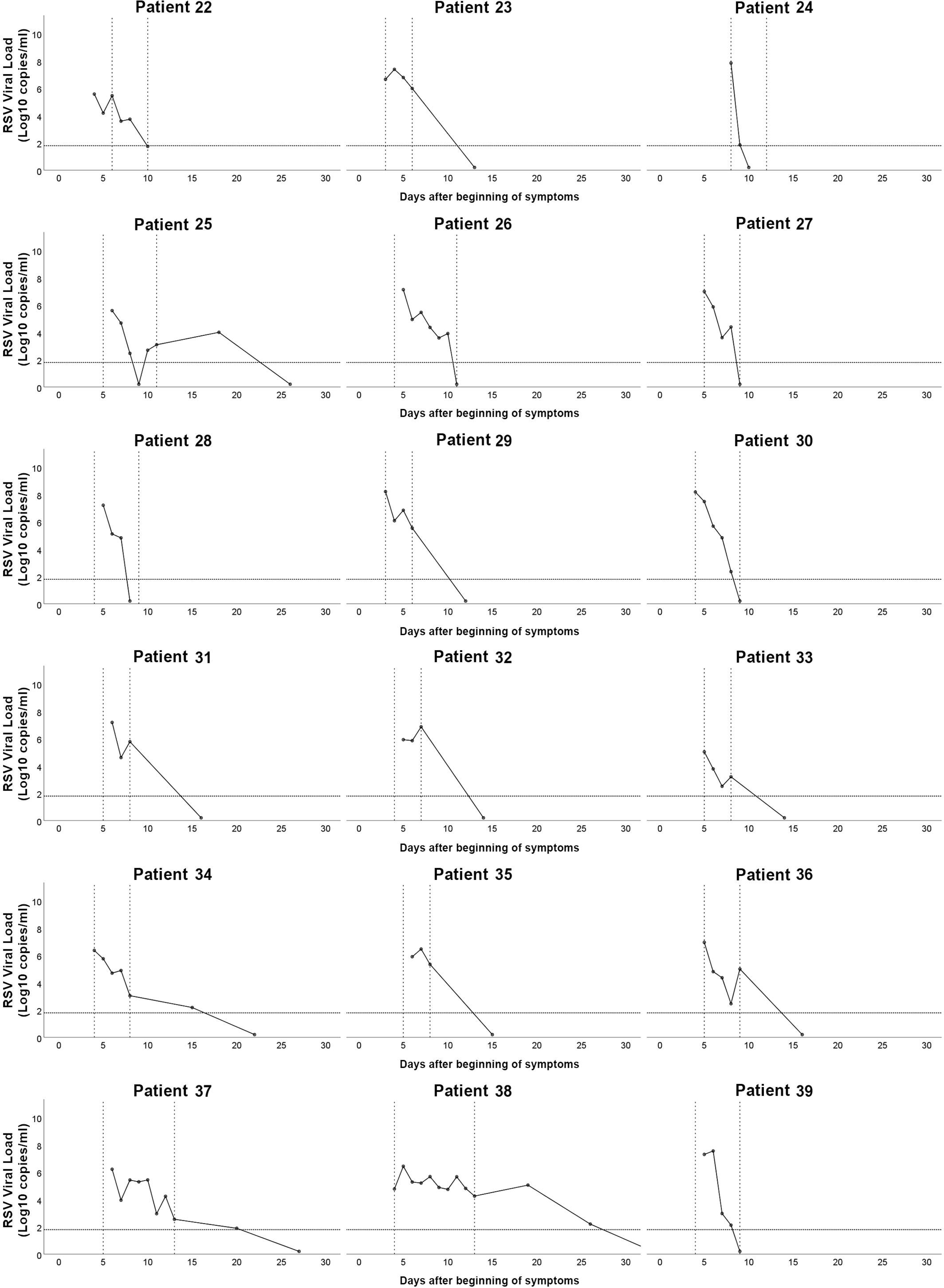
**RSV viral load decay, patients 22 to 39. Horizontal line correspond to the cut of viral detection. Vertical lines indicate period of hospitalization.**

**Figure 3.**
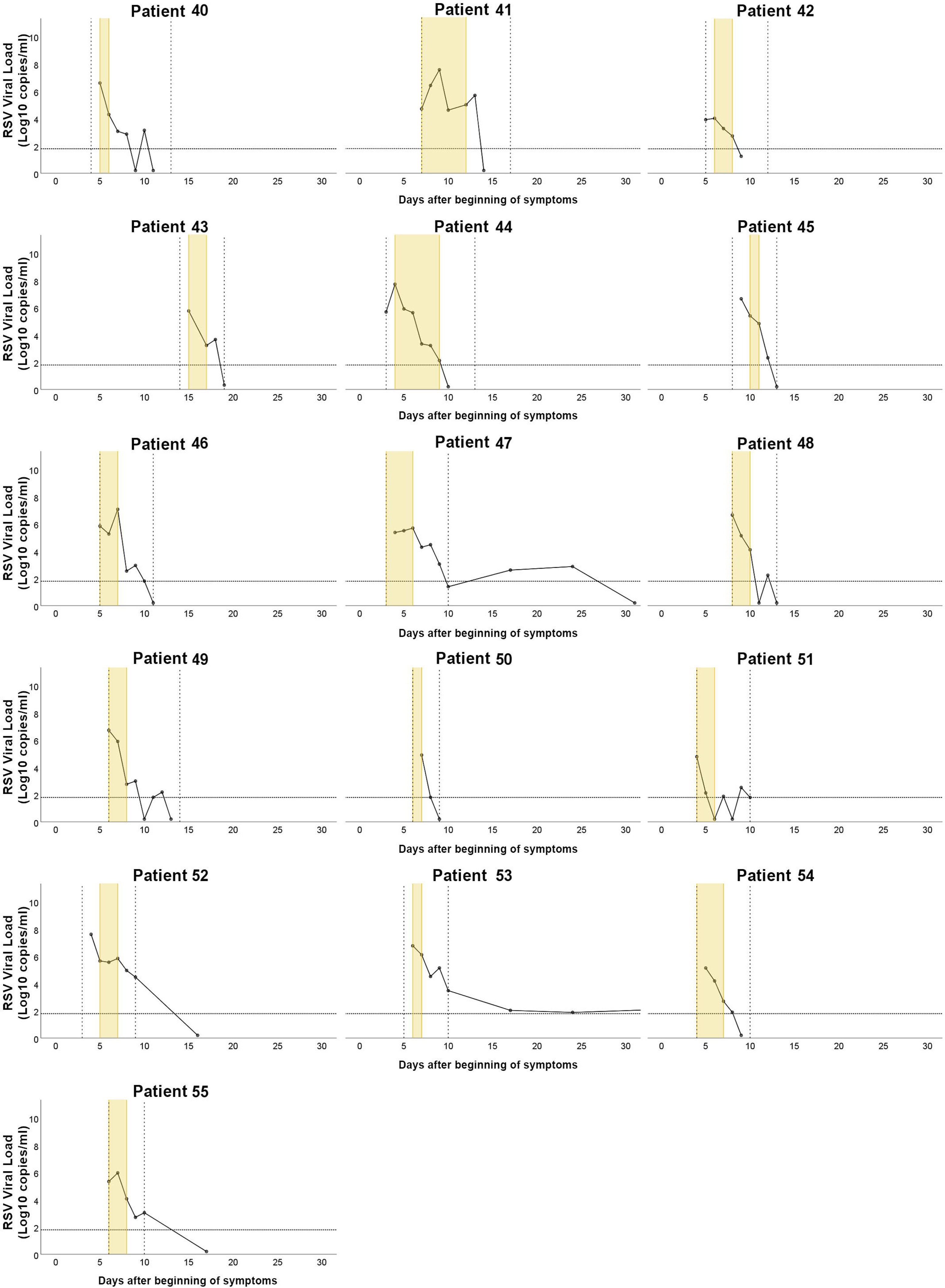
**RSV viral load decay, patients 40 to 55. Horizontal line corresponds to the cut of viral detection. Vertical lines indicate period of hospitalization. Yellow strips correspond to intensive care unit period.**

**Figure 4.**
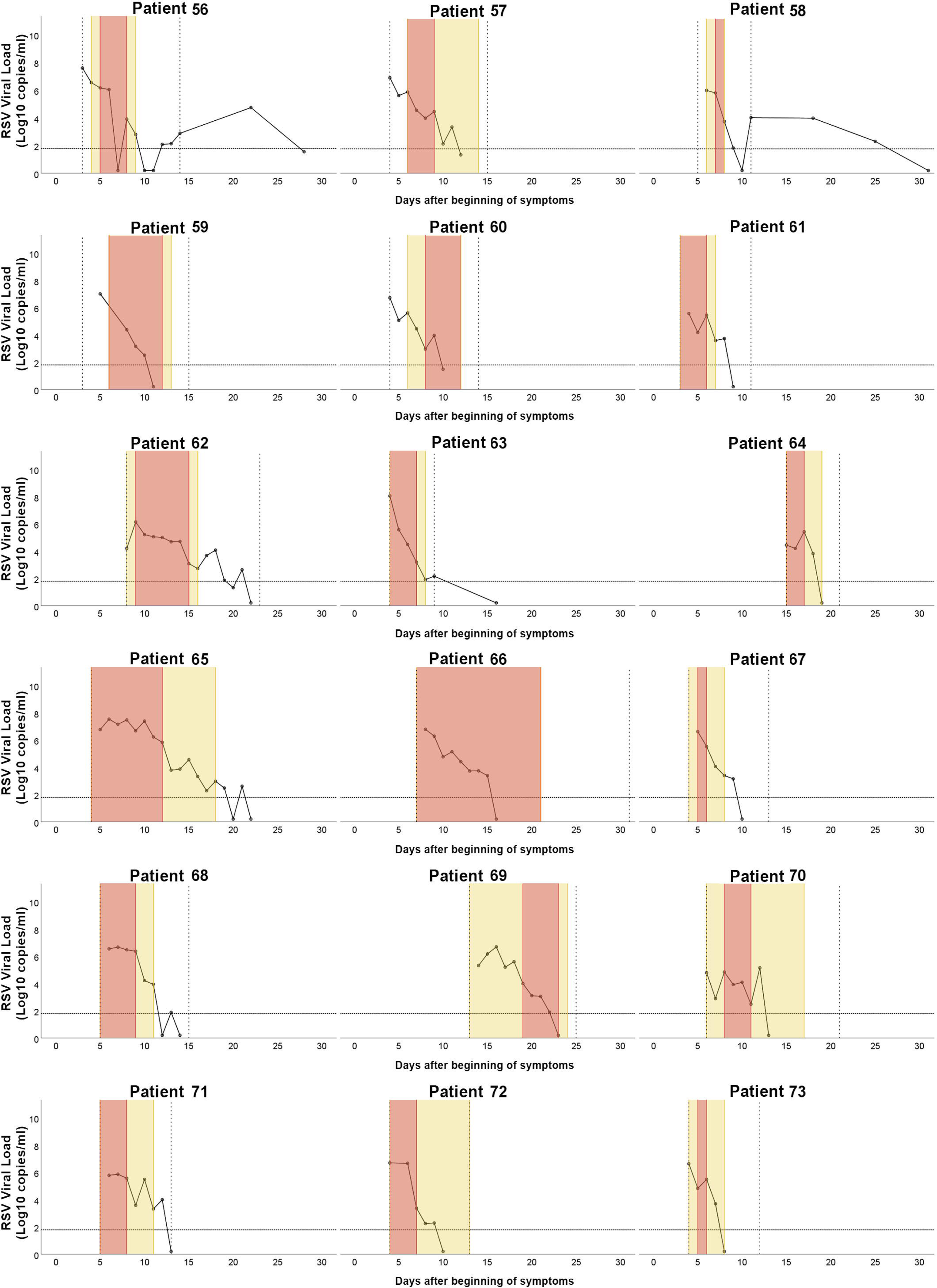
**RSV viral load decay, patients 56 to 73. Horizontal line corresponds to the cut of viral detection. Vertical lines indicate period of hospitalization. Yellow strip correspond to intensive care unit period and red strip to invasive mechanical ventilation period.**

**Figure 5.**
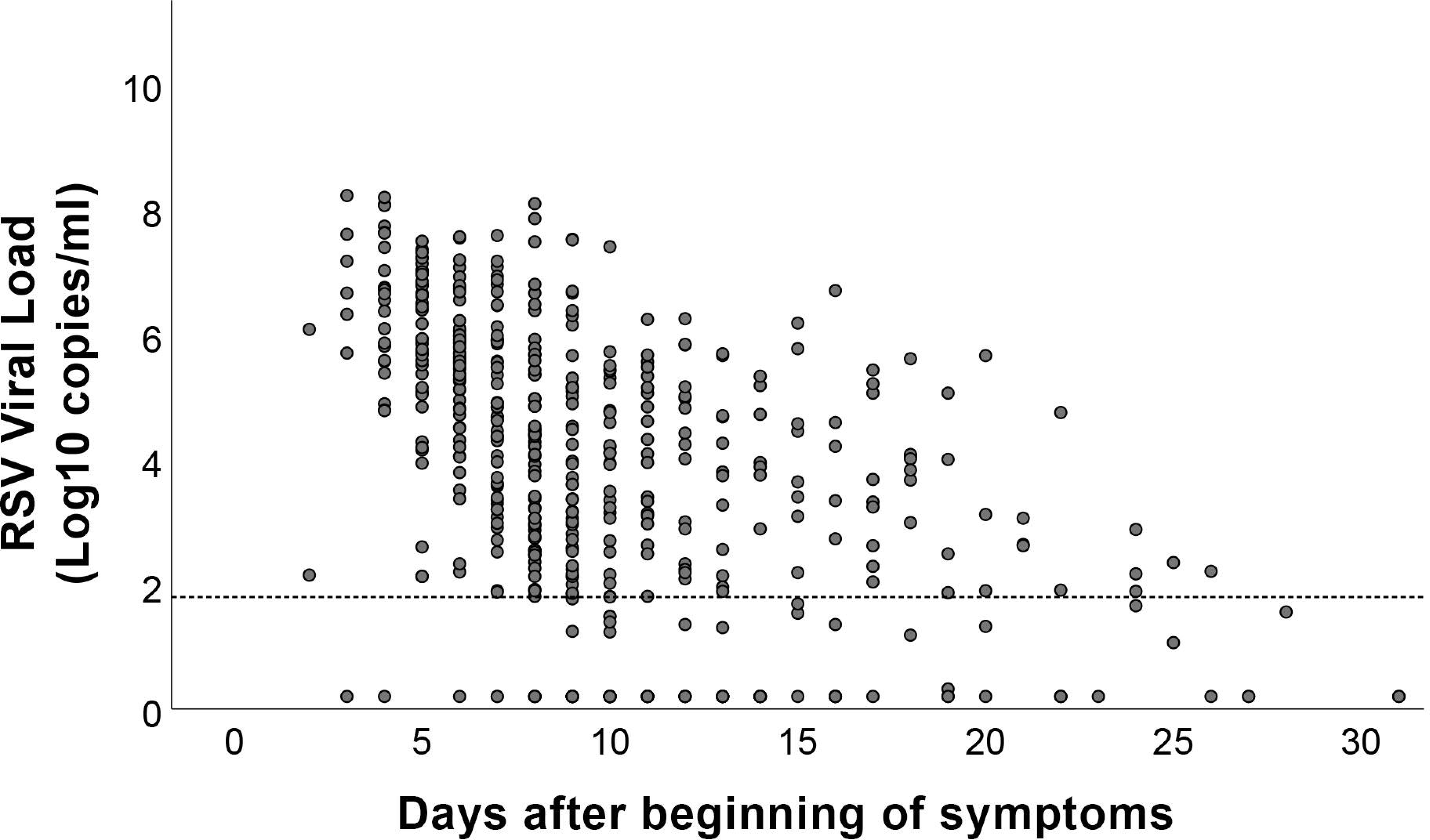
**RSV viral load of samples from all patients according to symptom onset.**

The duration of viral shedding (viral persistence) was determined as the last day of RSV detection in relation to symptom onset. The mean viral persistence was 12.5 ± 5.72 days, with a median of 10 days (ranging from 9 to 15 days) and varying from 6 to 33 days. A total of 33 patients (45.2%) exhibited viral persistence exceeding the median of 10 days. To provide a more detailed perspective, viral persistence was further analyzed on a weekly basis, considering that samples were collected every 7 days, as displayed in Table 3.

No statistically significant associations were found between viral load markers (first viral load, peak viral load, and viral persistence) and the various covariates, including age, gender, RSV subgroup, comorbidities, rhinovirus co-detection, and recurrent wheezing. The complete linear regression analysis for the model A is shown on table 4.

### Analysis of severity outcomes

Frequency and length of studied outcomes are described on table 5. Severity outcomes were studied as dependent variables of the multiple linear regression. Possible variables of association were analyzed as covariates. Age was inversely correlated with higher hospitalization period (p=0.039), more days of oxygen use (p=0.039) and need for ICU (p=0.045). Patients who experienced viral persistence for more than 10 days were found to have a significant correlation with a longer hospitalization period (p=0.009), increased days of ventilation support (p=0.044), extended duration of IMV (p=0.035), prolonged requirement for nutritional support (p=0.038), and a longer course of antibiotic treatment (p=0.024). Mean and standard deviation of studied outcomes and p value for patients according to the 10-day RSV shedding are show on table 6. The complete linear regression analysis for model B is shown on table 7.

## Discussion

Our study presents the most extensive longitudinal examination of RSV viral load in AVB patients, with an analysis based on severity outcomes. Despite utilizing a convenience sample over a two-year period, we successfully detected RSV in 64% of AVB patients that were submitted to viral detection. The study spanned two years, with a gap between them due to the outbreak of the SARS-CoV-2 pandemic, which prompted isolation measures and the closure of schools. This was justified by the fact that several other studies reported a delayed RSV outbreak in the first year of SARS-CoV-2 pandemic^22–24^. A previous study from our group described the patients from the pre-pandemic period^25^. Nevertheless, our findings align with the rates reported in existing literature^3,7^. It’s worth noting that the pandemic’s impact was evident in the second year of the study, with the absence of children experiencing recurrent wheezing.

Upon hospital admission, the reported time since symptom onset averaged 5 days, aligning with the point in the disease course described in the literature when symptoms typically worsen^7^.

Our study revealed several findings that were consistent with existing literature. These included a majority of male patients, a higher incidence of the disease among younger patients (particularly those under 6 months of age), and a predominance of RSV subgroup A^7,26–29^. Notably, the proportion of severe outcomes in our study, such as the need for ICU admission (46.6%) and IMV (24.7%), was significantly higher than previously reported. This could be attributed to the exclusion of patients who were discharged from the hospital early and the specific characteristics of our hospital^7,30^.

### Viral load dynamics

Viral load and viral persistence have been the subject of investigation in several studies aiming to comprehend RSV viral decay. For instance, Takeyama et al. and Hall et al. collected daily RSV samples during hospitalization but did not continue to monitor patients after discharge^9,31^. Takeyama et al. reported similar findings, including the time to reach peak viral load and the absence of an association between viral load and RSV subgroup with disease severity.

In the study conducted by Brint et al.^32^, RSV samples were collected daily throughout the hospitalization period and for specific days following discharge, extending up to the 28th day, even when RSV was no longer detectable. In that study, 11.8% of patients exhibited RSV detection for over 1 month, which was a higher rate compared to our findings. The discrepancy in results, including a higher rate of viral load rebound (19.6%) compared to our study (12.3%), could potentially be attributed to variations in viral load measurement techniques and the systematic collection of samples even after RSV was no longer detectable. Viral load rebound was defined as a 10-fold increase in the viral load after a nadir, which happened after the 8^th^ day of symptom onset, in two consecutive viral loads.

Viral load and viral persistence were not found to be associated with any of the patient characteristics (age, sex, comorbidities), RSV subgroup, or the presence of rhinovirus. This aligns with findings from other studies^9,32,33^.

### Severity Outcomes

Age was inversely associated with most severxity outcomes, as widely reported^3,28^. Our study could not find any association between quantitative viral load and severity outcomes. This association is controversial in the literature, with studies showing a positive association^8,11^, inverse association^12,33^ and others bringing no association at all^34,35^. Also, RSV subtype was not associated with severity outcomes; some studies corroborate this finding^36,37^, while others associate disease severity with the subgroup A^38,39^.

The primary and original data in our study was the association between viral persistence exceeding 10 days and most severity outcomes. The viral persistence may be a red flag for worsening outcomes and could encourage RSV detection and viral load quantification in patients with prolonged need for hospitalization and critical care. Future studies regarding treatment with antivirals and monoclonal antibodies could also address the benefit in RSV AVB patients with prolonged viral shedding.

Several limitations were experienced in our study. The accuracy of caregiver-provided information regarding the onset of symptoms, essential for standardizing the viral load curve, may be influenced by various factors, particularly prior upper airway infections, especially given the seasonal nature of respiratory viruses. There was an important loss of patients due to hospital transfer, as our hospital ward was not sufficient to hold all the patients during the peak of respiratory viruses’ seasonality. We also expected more patients with comorbidities, and for that reason we could not analyze them separately. Additionally, we did not explore the co-detection of other viruses and RSV genotypes, apart from the subgroup. It’s important to note that the severity of AVB can be influenced by a multitude of factors related to the virus, the patient, and external variables. Investigating all these factors simultaneously is a challenging in study design.

## Conclusion

Predictors of bronchiolitis severity can be highly complex. In addition to younger age, our study suggests that RSV viral persistence is associated to more severe disease. Further research into the dynamics of viral load in new studies may validate these findings. The information we have provided on viral shedding can contribute to a better understanding of RSV disease, particularly as several interventions, including vaccines, antivirals, and monoclonal antibodies, are anticipated to become available in the coming years.

## Data Availability

All data produced in the present study are available upon reasonable request to the authors

## Abbreviations

ICU: Intensive care unit
IMV: Invasive mechanical ventilation
RSV: Respiratory syncytial virus
RT-qPCR: Reverse Transcription - quantitative Polymerase Chain Reaction
SARS-CoV-2: Severe acute respiratory syndrome coronavirus 2

## Contributors Statement Page

**Raí André Silva Watanabe:** data curation, formal analysis, investigation, project administration, visualization, writing - original draft.

**Luciano de Souza Luna:** investigation, methodology, validation.

**Danielle Dias Conte, Ana Helena Perosa:** investigation.

**Larissa Lyra, Filipe Nishiyama, and Bruno Sanchez Camargo:** investigation, formal analysis.

**Nancy Cristina Junqueira Bellei:** conceptualization, funding acquisition, supervision, writing - review and editing.

## Ethics statement

Ethics committee of UNIVERSIDADE FEDERAL DE SAO PAULO gave ethical approval for this work under N. 3.009.916 - CEP/UNIFESP n. 1082/2018. Consent was given from participants legal representatives to take part in the present study.

