## Supplementary material for "Prolonged viral shedding as a marker of severity in respiratory syncytial virus bronchiolitis": table 1

Table 1. Characteristics of study population

| **Characteristics** | **Frequency** |
| --- | --- |
| **Sex** |  |
| Male | 44 (60.3%) |
| Female | 29 (39.7%) |
| **Age (months)** |  |
| Mean (SD) | 5.76 (5.66) |
| Median (IQR) | 4.3 (1.7 – 7.4) |
| **Comorbidities** |  |
| Present | 15 (20.5%) |
| Absent | 58 (79.5%) |
| **Recurrent wheezing** |  |
| Present | 8 (11.0%) |
| Absent | 65 (89.0%) |
| **VSR Subgroup** |  |
| A | 38 (52.1%) |
| B | 22 (30.1%) |
| Indetermined | 13 (17.8%) |
| **Other viruses** |  |
| Rhinovirus | 12 (16.4%) |
| Influenza | 0 (0%) |
