## Supplementary material for "Prolonged viral shedding as a marker of severity in respiratory syncytial virus bronchiolitis": table 2

Table 2. Time to reach peak viral load according to symptom onset

|  | **Day after symptom onset** | **Number of patients** | **Proportion of total** | **Accumulated proportion** |
| --- | --- | --- | --- | --- |
|  | 3rd day | 2 | 2.7% | 2.7% |
|  | 4th day | 16 | 21.9% | 24.7% |
|  | 5th day | 15 | 20.5% | 45.2% |
|  | 6th day | 13 | 17.8% | 63% |
|  | 7th day | 10 | 13.7% | 76.7% |
|  | 8th day | 4 | 5.5% | 82.2% |
|  | 9th day | 6 | 8.2% | 90.4% |
|  | After 10 days | 7 | 9.6% | 100% |
