## Supplementary material for "Prolonged viral shedding as a marker of severity in respiratory syncytial virus bronchiolitis": table 3

Table 3. Weekly viral shedding from patients according to symptom onset

| **Viral shedding** | **Number of Patients** | **Proportion of total** | **Accumulated percentage** |
| --- | --- | --- | --- |
| Up to 7 days | 7 | 9.6% | 9.6% |
| 8 to 14 days | 47 | 64.4% | 74% |
| 15 to 21 days | 12 | 16.4% | 90.4% |
| 22 to 28 days | 6 | 8.2% | 98.6% |
| Over 28 days | 1 | 1.4% | 100% |
