## Supplementary material for "Prolonged viral shedding as a marker of severity in respiratory syncytial virus bronchiolitis": table 4

**Table 4: Multiple linear regression for model A**

| **Covariates** | | **Dependent variables** | | |
| --- | --- | --- | --- | --- |
|  |  | **Viral persistence** | **First viral load** | **Peak viral load** |
| **Age** | **β1** | 0.599 | -0.053 | -0.290 |
|  | **p** | 0.671 | 0.051 | 0.420 |
| **Sex**  **(Male – Female)** | **β2** | 2.338 | 0.369 | 0.121 |
|  | **p** | 0.992 | 0.243 | 0.922 |
| **RSV Subtype**  **(A – B)** | **β3** | -0.206 | 0.249 | 0.068 |
|  | **p** | 0.904 | 0. 438 | 0.202 |
| **Comorbidities*** | **β4** | -1.352 | 0.319 | 0.526 |
|  | **p** | 0.526 | 0.428 | 0.553 |
| **Rhinovirus*** | **β5** | 2.270 | --0.560 | -0.527 |
|  | **p** | 0.278 | 0.157 | 0.120 |
| **Intercept (β0)** | | 10.827 | 5.881 | 6.293 |
| **R^2^** | | 0.087 | 0.152 | 0.119 |

β1 to β5 = angular coefficient

RSV = Respiratory syncytial virus

* The reference level for the covariates is its absence
