## Supplementary material for "Prolonged viral shedding as a marker of severity in respiratory syncytial virus bronchiolitis": table 5

**Table 5.** **Frequency and length (days) of studied outcomes**

| **Outcome** | **Frequency** | **Mean (SD) *** | **Median (IQR) *** |
| --- | --- | --- | --- |
| **Hospitalization** |  | 8.12 (3.99) | 7 (6 - 10) |
| **Fever** | 41 (56.2%) | 2.27 (1.14) | 2 (1 – 3) |
| **Oxygen** | 72 (98.6%) | 5.61 (3.37) | 5 (3.75 – 7) |
| **ICU** | 34 (46.6%) | 5.82 (3.59) | 5 (3 – 7) |
| **Any ventilation support** | 32 (43.8%) | 4.84 (3.32) | 4 (2.75 – 6.25) |
| **IMV** | 18 (24.7%) | 4.94 (2.90) | 4 (4 – 5) |
| **Nutrition support** | 34 (46.6%) | 6.38 (3.98) | 6 (4 – 7) |

* Values only from patients that presented each of the outcomes, not from the total
