## Supplementary material for "Prolonged viral shedding as a marker of severity in respiratory syncytial virus bronchiolitis": table 6

**Table 6.** **Mean and standard deviation (days) of studied outcomes and *p* value for patients according to the 10-day RSV shedding**

| **Outcome** | **RSV shedding**  **over 10 days** | **Mean (SD)** | *p* value |
| --- | --- | --- | --- |
| **Hospitalization** | Yes | 9.7 (4.9) | 0.002 |
|  | No | 6.8 (2.4) |  |
| **Fever** | Yes | 1.5 (1.5) | 0.326 |
|  | No | 1.1 (1.4) |  |
| **Oxygen** | Yes | 6.3 (4.3) | 0.080 |
|  | No | 4.9 (2.3) |  |
| **ICU** | Yes | 3.7 (4.7) | 0.037 |
|  | No | 1.9 (2.7) |  |
| **Any ventilation** | Yes | 3.0 (4.0) | 0.043 |
|  | No | 1.4 (2.3) |  |
| **IMV** | Yes | 1.8 (3.3) | 0.057 |
|  | No | 0.7 (1.7) |  |
| **Nutrition support** | Yes | 4.1 (5.2) | 0.032 |
|  | No | 2.0 (2.9) |  |
| **Antibiotics** | Yes | 4.7 (5.0) | 0.011 |
|  | No | 2.2 (3.0) |  |
