## Supplementary material for "Prolonged viral shedding as a marker of severity in respiratory syncytial virus bronchiolitis": table 7

**Table 7: Multiple linear regression for model B**

| **Covariates** | | **Outcomes/Dependent variables (days)** | | | | | | | |
| --- | --- | --- | --- | --- | --- | --- | --- | --- | --- |
|  |  | **Hospitalization** | **Fever** | **Oxygen** | **ICU** | **Any Ventilation** | **IMV** | **Nutrition** | **Antibiotics** |
| **Age** | **β1** | **- 0.007** | 0.060 | **-0.141** | **-0.191** | -0.124 | -0.115 | -0.145 | -0.114 |
|  | **p** | **0.039** | 0.100 | **0.039** | **0.045** | 0.129 | 0.072 | 0.170 | 0.276 |
| **Peak viral load** | **β2** | 0.046 | 0.079 | 0.145 | -0.219 | -0.128 | 0.013 | -0.377 | -0.048 |
|  | **p** | 0.936 | 0.713 | 0.778 | 0.695 | 0.792 | 1.000 | 0.548 | 0.938 |
| **Viral persistence over 10 days*** | **β3** | **3.006** | 0.513 | 1.919 | 2.182 | **1.964** | **1.598** | **2.616** | **2.826** |
|  | **p** | **0.009** | 0.230 | 0.062 | 0.052 | **0.044** | **0.035** | **0.038** | **0.024** |
| **Sex**  **(Male – female)** | **β4** | 0.114 | 0.032 | -0.102 | -0.126 | -0.232 | -0.243 | -0.171 | 0.692 |
|  | **p** | 0.992 | 0.942 | 0.922 | 0.912 | 0.814 | 0.751 | 0.893 | 0.583 |
| **RSV subtype**  **(A – B)** | **β5** | -0.268 | 0.282 | 0.537 | 0.795 | 0.653 | 0.318 | 0.951 | -0.491 |
|  | **p** | 0.816 | 0.513 | 0.602 | 0.479 | 0.503 | 0.675 | 0.451 | 0.693 |
| **Comorbidities*** | **β6** | 1.220 | -0.165 | 0.526 | -0.721 | -0.663 | 0.563 | -0.376 | 1.467 |
|  | **p** | 0.389 | 0.755 | 0.676 | 0.600 | 0.578 | 0.544 | 0.807 | 0.337 |
| **Rhinovirus*** | **β7** | -0.157 | -0.178 | -0.732 | -1.271 | -1.278 | -0.969 | -1817 | -0.753 |
|  | **p** | 0.911 | 0.737 | 0.562 | 0.356 | 0.285 | 0.298 | 0.241 | 0.621 |
| **Intercept (β0)** | | 7.739 | 0.116 | 4.486 | 4.409 | 3.004 | 1.272 | 5.195 | 2.997 |
| **R^2^**  **p** | | 0.226  0.004 | 0.070 | 0.142 | 0.195 | 0.167 | 0.177 | 0.163 | 0.161 |

β1 a β7 = angular coefficient

RSV = Respiratory syncytial vírus

ICU = Intensive care unit

IMV = Invasive mechanical ventilation

* The reference level for the covariates is its absence
